# Microplastics Identified in Commercial Over-the-counter Lubricant Eyedrops

**DOI:** 10.1101/2024.05.26.24307941

**Authors:** Julia E Jaeger, Matthew Burke, Duoduo Wu, Emily Ern-Min Curren, Sandric Chee Yew Leong, Robert Symons, Blanche Xiao Hong Lim, Xinyi Su, Jodhbir Singh Mehta, Andri Kartasasmita Riau, Chris Hong Long Lim

## Abstract

**Purpose:** There is increasing evidence that microplastics exert harmful effects on human health and on the ocular surface. Recent studies have highlighted the presence of ocular surface exposure to microplastics via shedding from contact lenses. This study aims to investigate if microplastic particles are present in commonly used eyedrops in single-use plastic vials.

**Design:** Experimental study.

**Methods:** Nine commonly used commercial tear-replacement solutions available without a doctor’s prescription were tested (Brands A-I). All brands of eyedrops were analysed visually using light microscopy and the number of microplastic particles were manually counted. Brands A-F were further analysed using the 8700 Agilent Laser Direct Infrared (LDIR) chemical imaging system to identify the microplastic compositions.

**Results:** All eyedrops analysed contained microplastics. The number of microplastic particles identified using light microscopy ranged between 15 (Brand E) to >18,000 (Brand F). In total, nine types of microplastics were identified with LDIR – an average of 14 polyethylene, 8 polypropylene, 1 polystyrene, 2 polyvinylchloride, 21 polyethylene terephthalate, 1 polycarbonate, 19 polymethylmethacrylate, 23 polyamide and 22 polyurethane per millilitre of eyedrop were identified.

**Conclusions:** This is the first study that identified microplastics in commercial tear-replacement solutions. These particles may have been derived from secondary degradation of the plastic vials (polyethylene) during production, transportation, or storage, and represent a major source of exposure to the ocular surface, especially among patients who require chronic instillation of eyedrops.

## Introduction

Plastics are synthetic or semi-synthetic polymeric materials found commonly in everyday life. Their plasticity allows them to be moulded into objects with varied shapes and rigidity, making them invaluable materials in daily life and especially in healthcare ^1^. Unfortunately, plastics have been shown to undergo secondary degradation into smaller pieces, termed microplastics and nanoplastics, when environmental factors such as mechanical abrasion, ultraviolet irradiation and high temperatures are applied.

Microplastics are defined as plastic particles smaller than 5 mm ^2^. Primary microplastics are manufactured plastic particles of small sizes, while secondary microplastics are derived from the physical, chemical, and biological degradation of larger plastic materials found in the environment. Their presence is ubiquitous in daily living, and studies have identified microplastics in air, water, food, and humans ^3,4^. As a sector heavily reliant on plastics for its low cost, sterility, and plasticity, microplastics have been detected in the healthcare surgical environment ^5^. Worryingly, microplastics were recently isolated from 49 vitreous samples of patients who underwent intraocular surgery at two centres in China ^6^.

There is increasing evidence that microplastics exposure may pose health risks and has been implicated in the development of malignancies, endocrine and hormonal abnormalities, and abnormal foetal development ^4,7–9^. Exposure of the ocular surface to microplastics is associated with upregulation of IL-1α, IL1-β, and IL-6 in the conjunctiva and lacrimal glands, as demonstrated in murine models ^10^. Microplastics are also present in the atmosphere as particulate matter, and population-based studies have suggested that high levels of these particles may be associated with the development of dry eye disease ^11–13^. Elevated concentrations of microplastics in vitreous samples were also associated with higher intraocular pressures and the presence of aqueous humour opacities ^6^. Contact lenses have been identified as a source of microplastics, with an estimated release of 90,000 particles on the ocular surface from regular wear of ten hours daily over a year ^14^. Given the ubiquitous nature of microplastics in plastic packaging, it is conceivable that microplastics may also be present in commercially available eyedrops. This is of particular concern as patients with chronic ophthalmic conditions such as glaucoma, dry eye disease or uveitides may require long-term instillation of eyedrops which exposes the ocular surface to microplastics. Thus far, there is no published literature supporting the identification of microplastics in topical ophthalmic formulations. This study aims to identify and characterise microplastics in common over-the-counter tear-replacement solutions to ascertain whether there is a potential for microplastic exposure of the ocular surface when eye drops are administered.

## Methodology

### Sample Collection

Single-use tear-replacement solutions of various brands and formulations were selected for this study. Over-the-counter commercial eyedrops with their constituent ingredients and volumes are summarised in Table 1. These formulations were chosen as they are commonly prescribed lubricants and can also be bought off-the-shelf from departmental stores and retail pharmacies without a prescription.

**Table 1:** Composition of selected tear-replacement solutions.

| Brand | Ingredients | Volume |
| --- | --- | --- |
| A | Carmellose sodium*<br>Sodium chloride<br>Sodium lactate<br>Potassium chloride<br>Calcium chloride dihydrate<br>Magnesium chloride hexahydrate<br>Purified water<br>Sodium hydroxide<br>Hydrochloric acid | 0.4 ml |
| B | Carboxymethylcellulose sodium*<br>Sodium chloride<br>Sodium lactate<br>Potassium chloride<br>Calcium chloride dihydrate<br>Magnesium chloride hexahydrate<br>Purified water<br>Sodium hydroxide<br>Hydrochloric acid | 0.4 ml |
| C | Carboxymethylcellulose sodium*<br>Glycerin*<br>Boric acid<br>Calcium chloride dihydrate<br>Erythritol<br>Levocarnitine<br>Magnesium chloride hexahydrate<br>Potassium chloride<br>Purified water<br>Stabilized oxychloro complex<br>Sodium borate decahydrate<br>Sodium citrate dihydrate | 0.4 ml |
| D | Polyvinyl Alcohol*<br>Povidone*<br>Sodium chloride<br>Purified water<br>Sodium hydroxide<br>Hydrochloric acid | 0.4 ml |
| E | Dextran*<br>Hypromellose*<br>Potassium chloride<br>Purified water<br>Sodium borate<br>Sodium chloride<br>Sodium hydroxide<br>Hydrochloric acid | 0.8 ml |
| F | Carboxymethylcellulose sodium*<br>Glycerin*<br>Polysorbate*<br>Boric acid<br>Carbomer copolymer type A<br>Castor oil<br>Erythritol<br>Levocarnitine<br>Purified water<br>Sodium hydroxide<br>Hydrochloric acid | 0.4 ml |
| G | Polyethylene glycol*<br>Propylene glycol*<br>Hydroxypropyl guar<br>Sorbitol<br>Aminomethyl Propanol<br>Boric acid<br>Potassium chloride<br>Sodium chloride | 0.7 ml |
| H | Polyethylene glycol 400*<br>Sodium chlorite<br>Sodium hyaluronate<br>Boric Acid<br>Sodium borate<br>Calcium chloride (dihydrate)<br>Magnesium Chloride<br>Sodium chloride<br>Potassium chloride | 0.4 ml |
| I | Sodium carboxymethylcellulose*<br>Sodium hyaluronate*<br>Glycerin<br>Erythrol<br>Levocarnitine<br>Sodium lactate<br>Calcium chloride dihydrate<br>Magnesium chloride hexahydrate<br>Potassium chloride | 0.4 ml |

\*Active ingredient(s)

All eyedrop samples were analysed prior to their expiry date. Two methods of analyses were utilised in a microplastic-free environment – light microscopy with manual count and laser direct infrared (LDIR) imaging system (Figure 1). Details of the analyses are outlined below.

**Figure 1:**
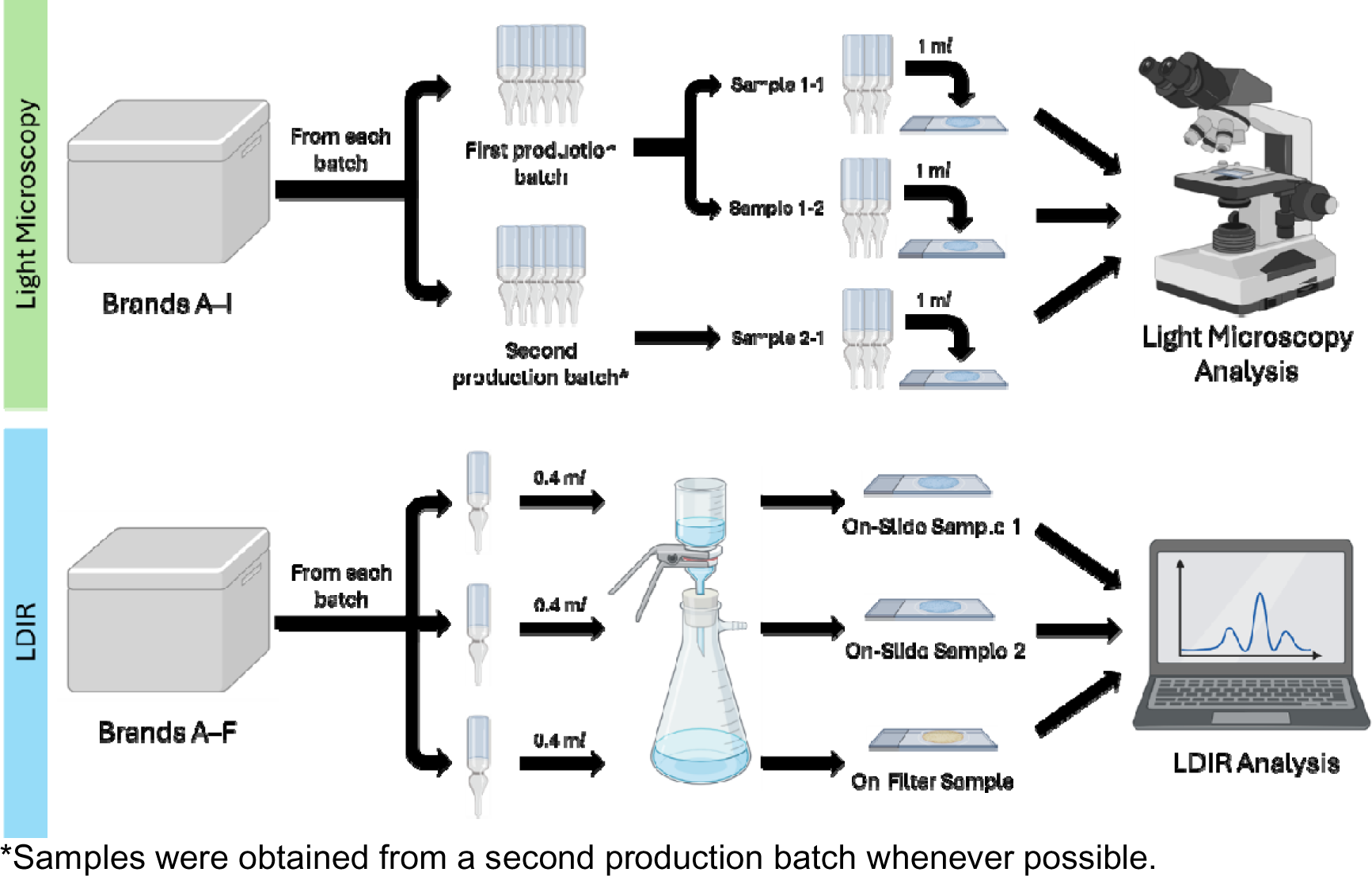
Summary experimental workflow of analysis with light microscopy and laser direct infrared imaging system with spectroscopy.

### Light Microscopy

Prior to analysis, all labware including forceps, glass bottles, petri dishes and metal sieves were sterilised with ethanol and rinsed with Mili-Q ultrapure water twice. In addition, glass and stainless steel labware were used to minimise external microplastic contamination. All open-lid containers were covered with either a glass slide or aluminium foil to minimise microplastic contamination from the air. All loading of eyedrop samples were conducted in a clean air cabinet. Contamination protocols undertaken during analysis are as described by Curren and Leong ^15^.

A total of three samples of each over-the-counter tear-replacement solution were obtained. Two samples were from the same production batch while a third sample was obtained from another batch when possible. This allowed for comparison of microplastic counts between samples of the same batch and between batches for each formulation, to determine the consistency of microplastic counts between batches. Samples from tear-replacement solutions were analysed by pipetting 1 mL of each eyedrop formulation using a glass pipette onto a Sedgewick rafter slide and viewed under an inverted light microscope. Microplastic particles were identified based on visual characteristics and sorted into categories such as fibre, film, or fragment. The number of microplastics in each sample was determined from triplicate counts. A blank controls were used prior to each eyedrop samples count analysis to ensure that no ambient microplastic contamination was present prior to testing.

### Laser Direct Infrared Imaging System with Spectroscopy

LDIR is an infrared spectrometer that utilises a fast-tuneable quantum-cascade laser (QCL) coupled with a rapidly scanning imaging system to analyse particulate information such as quantification, size, colour and morphology, while identification of polymer type can be identified with spectroscopic analysis ^16^.

To prevent environmental contamination, the LDIR analysis was performed in a segregated room within the facility, physically distinct and equipped with an airlock system and change pod. The laboratory was kept at a positive pressure and air inflow HEPA filtered. Lab analysts also don specialised black cotton lab coats with efforts made to avoid clothing that is prone to shedding polymer fibres. All glassware underwent either furnace cleaning for a minimum of 4 hours at temperatures exceeding 400 °C or washing with microplastics-analysis-grade (MAG) water. Prior to use, MAG water samples were analysed to ascertain their purity. Sample filtration was performed within a fume cupboard, with samples left in the cupboard for over 5 minutes covered with foil. Monthly air blank tests are conducted to assess the air quality within the laboratory and the fume cupboard. Nightly vacuuming and regular surface wiping are also conducted to remove potential contaminants. Quality control and accuracy of analysis was assessed by running samples with a known concentration of green polyethylene (PE) and analysing reagent blanks using 1 L of MAG water. A polymer register is also diligently updated to track and trace potential contaminations.

Reference materials for the analysis were obtained from various manufacturers: fluorescent green PE microspheres of 75-90 µm and 250-300 µm (Cospheric, Santa Barbara, CA, USA), polypropylene (PP) chromatographic grade of 150 µm (Polysciences Inc, Warrington, PA, USA), polystyrene (PS) EZY-CALTM microsphere size standard, NIST-traceable with mean diameter 30 or 70 µm, and a concentration of 2,000 particles/mL (ThermoFisher Scientific Waltham, MA, USA), polyvinylchloride (PVC) analytical standard (Sigma-Aldrich, Saint Louis, MO, USA), polyethylene terephthalate (PET) with a maximum particle size of 300 µm (Goodfellow, Cambridge, United Kingdom), polycarbonate (PC) beads sized 150-250 µm (Sigma-Aldrich, Saint Louis, MO, USA), polymethylmethacrylate (PMMA) analytical standard with a size of 50 μm in solution (Sigma-Aldrich, Saint Louis, MO, USA), polyamide (PA6, Nylon) with a mean size of 15-20 µm (Goodfellow, Cambridge, United Kingdom), polyurethane (PU) 3-5 mm (Goodfellow, Cambridge, United Kingdom), and POLYMER KIT 1.0 (Hawaii Pacific University, Honolulu, HI, USA). MAG water was obtained by filtering ultrapure water from Arium® Mini water purification system connected to a CellPlus Ultrafilter (Sartorius, Göttingen, Germany) and three subsequent filtrations through a 5 μm polycarbonate filter.

To analyse microplastic content in commercially available tear-replacement solutions, two different kinds of analysis were used: MirrIR low-e microscope slides (Kevley Technologies, Chesterland, OH, USA) analysis (slide) and on-filter analysis (filter). A separate production batch of tear-replacement solutions was obtained for this analysis. For every brand of eyedrop, two replicates of on-slide analysis and one on-filter analysis were conducted. For slide analysis, the entire content (0.4 mL) of a tear-replacement vial was filtered through a 5 μm pore size polycarbonate filter paper (Sartorius, Göttingen, Germany) mounted in a glass vacuum filtration unit with a funnel (Rocker, Kaohsiung, Taiwan) attached to a vacuum pump (Wiggens, Wuppertal, Germany). The filter was then rinsed with 100 mL of MAG water. All particles were transferred onto a slide for analysis. On-filter analysis of the gold filter was also performed to avoid the potential loss due to the transfer. Filtration was performed with 25 mm diameter, 0.8 µm pore size gold-coated polycarbonate membrane filters (Sterlitech, Aubrun, WA, USA). The entire content (0.4 mL) of one eyedrop vial was administered through the filter directly and washed with 100 mL of MAG water. Once dry, the filter was mounted on a filter holder (Agilent Technologies, Santa Clara, CA, USA) and analysed. For the analysis of the tear-replacement vial compositions, small fragments < 300 µm of the vial have been cut off and placed onto a slide for analysis.

All samples were analysed to quantify polymer particles and size ranges between 20 and 500 µm on the Agilent 8700 LDIR chemical imaging system (Agilent Technologies, Santa Clara, CA, USA). Particles larger than 500 µm were not detected. The analyses were conducted by the automated Particle Analysis workflow as part of the Clarity software Version 1.5 (Agilent Technologies, Santa Clara, CA, USA). This workflow automatically counts and sizes all particles. High-magnification images can also be taken during the analysis. For particle identification, an IR spectrum is generated in the fingerprint region (1800 – 975 cm^−1^). The acquired spectra was then compared to an internal library. The library used in this study is based on siMPle, which has been modified for the use of the LDIR chemical imaging system ^17^. Additionally, information from open-source repositories such as μATR-FTIR Spectral Libraries of Plastic Particles (FLOPP and FLOPP-e) for the Analysis of Microplastics have been intergraded ^17,18^. Finally, the library was also modified by Eurofins Environment Testing for the nine reported polymers which were verified against two independently purchased reference materials.

The match quality was determined by comparing the acquired spectra to the library reference spectra. A higher quality match score indicates a better fit between the measured spectrum and the material’s spectrum, with a score of 1 representing an absolute match. For this study, a minimum match quality above 0.7 was deemed necessary.

## Results

The number of visually identified microplastic particles in each 1 mL sample via light microscopy is summarised with a logarithmic scale in Figure 2. Eyedrops from a separate production batch were not available for brands C, D, F and I, hence light microscopy analysis was performed only for two samples from within the same production batch. Notably, Brand F had a cloudy appearance during analysis, and yielded the largest number of possible microplastic particles (>18,000 per millilitre of eyedrop) among all the tear-replacement samples tested. Even among eyedrops from the same production batch, the number of microplastic particles remained varied.

**Figure 2:**
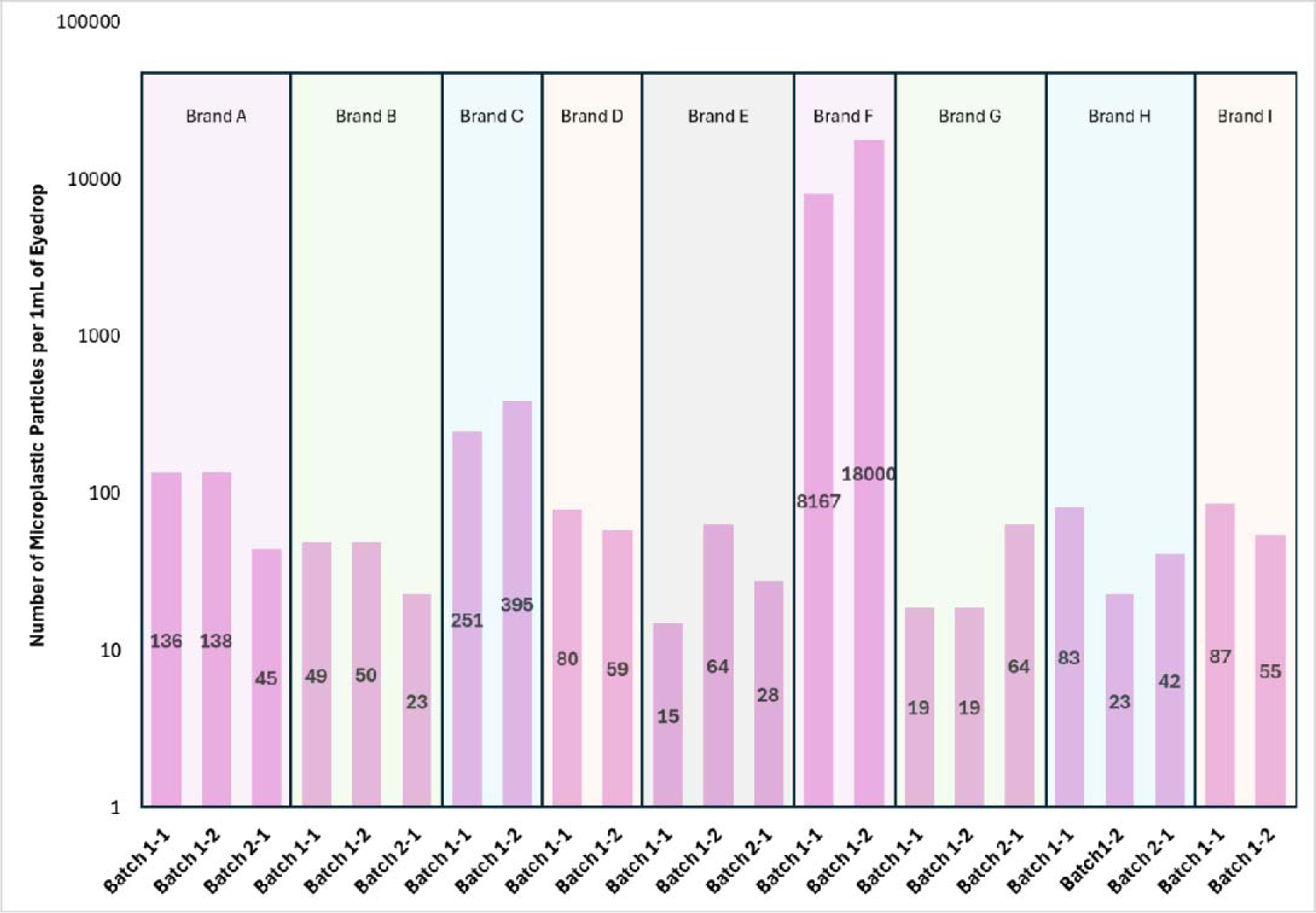
Number of visually identified microplastic particles using light microscopy. Samples were obtained from across two production batches from each tear-replacement brand when possible.

LDIR analysis was performed for seven tear-replacement brands (Brands A-F). Two replicates of on-slide analysis and one on-filter analysis were performed for all samples. All samples showed evidence of microplastic contamination (Table 2) (Figure 3). Nine types of microplastics were identified – polyamide (PA), polycarbonate (PC), polyethylene (PE), polyethylene terephthalate (PET), polymethylmethacrylate (PMMA), polypropylene (PP), polystyrene (PS), polyurethane (PU), and polyvinylchloride (PVC) (Figure 4). These microplastics were identified in varying levels in each tear-replacement sample and ranged between 20-500 μm. The amount and type of microplastic particles retrieved from each unit dose vial varied. This ranged between five and 308 particles per 1 mL of eyedrop (Table 2). The most identified microplastic types were PET, PA and PU, which were identified in 16, 15 and 14 samples respectively. Analysis of tear-replacement solution vials shows that they are made of PE.

**Figure 3:**
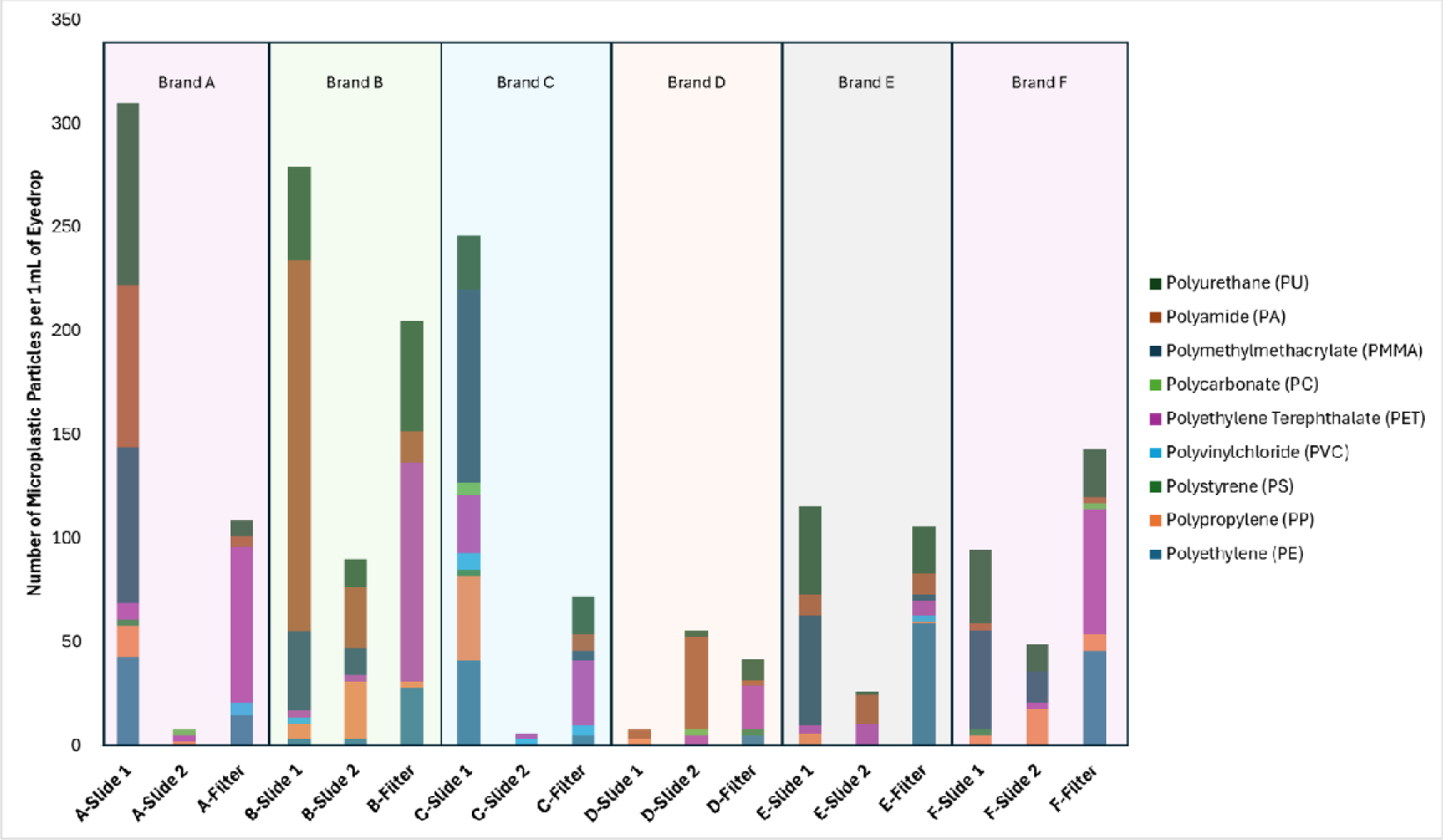
Number and type of microplastic particles identified using laser direct infrared imaging system with spectroscopy. Two different kinds of analyses were used: Kevley slide analysis (slide) and on-filter analysis (filter).

**Figure 4:**
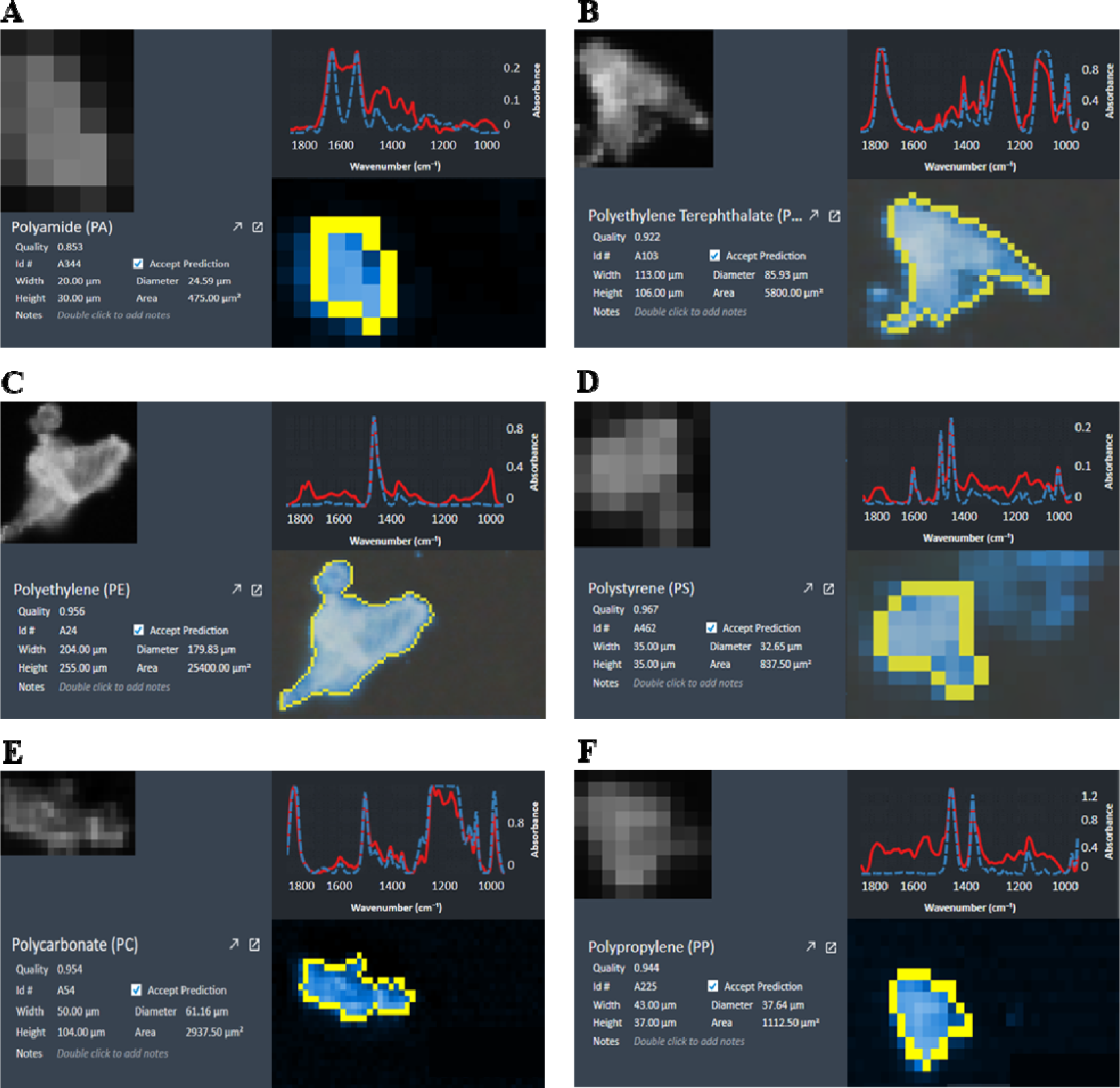
Representative images of (A) polyamide, (B) polyethylene terephthalate, (C) polyethylene, (D) polystyrene, (E) polycarbonate, and (F) polypropylene identified in tear-replacement solutions using laser direct infrared imaging system with spectroscopy. Blue dotted lines represent reference spectra and red solid lines represent measured spectra.

**Table 2:**
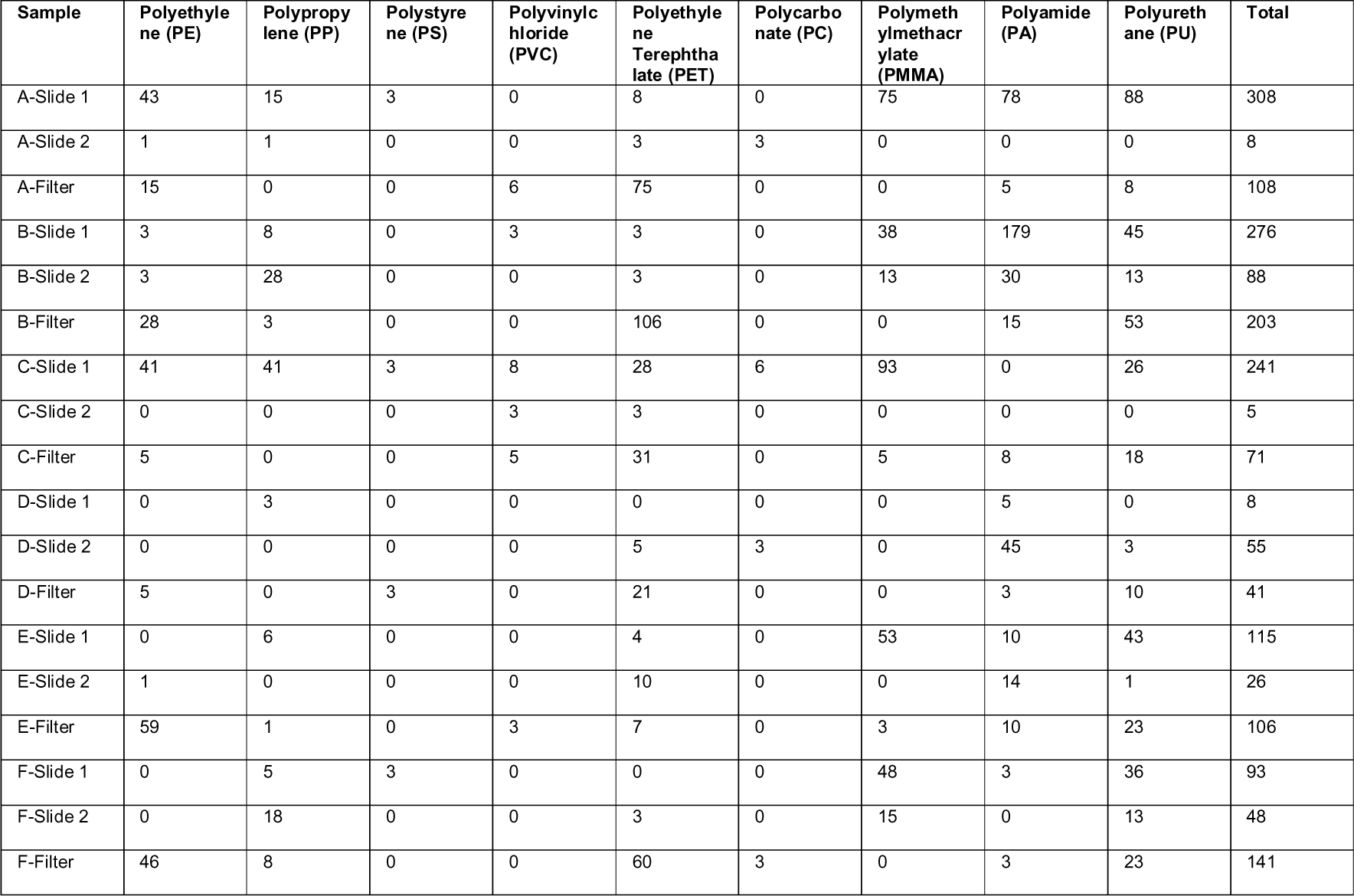
Number and type of microplastics per millilitre identified via laser direct infrared imaging system with spectroscopy.

| Sample | Polyethylene (PE) | Polypropylene (PP) | Polystyrene (PS) | Polyvinyl chloride (PVC) | Polyethylene Terephthalate (PET) | Polycarbonate (PC) | Polymethylmethacrylate (PMMA) | Polyamide (PA) | Polyurethane (PU) | Total |
| --- | --- | --- | --- | --- | --- | --- | --- | --- | --- | --- |
| A-Slide 1 | 43 | 15 | 3 | 0 | 8 | 0 | 75 | 78 | 88 | 308 |
| A-Slide 2 | 1 | 1 | 0 | 0 | 3 | 3 | 0 | 0 | 0 | 8 |
| A-Filter | 15 | 0 | 0 | 6 | 75 | 0 | 0 | 5 | 8 | 108 |
| B-Slide 1 | 3 | 8 | 0 | 3 | 3 | 0 | 38 | 179 | 45 | 276 |
| B-Slide 2 | 3 | 28 | 0 | 0 | 3 | 0 | 13 | 30 | 13 | 88 |
| B-Filter | 28 | 3 | 0 | 0 | 106 | 0 | 0 | 15 | 53 | 203 |
| C-Slide 1 | 41 | 41 | 3 | 8 | 28 | 6 | 93 | 0 | 26 | 241 |
| C-Slide 2 | 0 | 0 | 0 | 3 | 3 | 0 | 0 | 0 | 0 | 5 |
| C-Filter | 5 | 0 | 0 | 5 | 31 | 0 | 5 | 8 | 18 | 71 |
| D-Slide 1 | 0 | 3 | 0 | 0 | 0 | 0 | 0 | 5 | 0 | 8 |
| D-Slide 2 | 0 | 0 | 0 | 0 | 5 | 3 | 0 | 45 | 3 | 55 |
| D-Filter | 5 | 0 | 3 | 0 | 21 | 0 | 0 | 3 | 10 | 41 |
| E-Slide 1 | 0 | 6 | 0 | 0 | 4 | 0 | 53 | 10 | 43 | 115 |
| E-Slide 2 | 1 | 0 | 0 | 0 | 10 | 0 | 0 | 14 | 1 | 26 |
| E-Filter | 59 | 1 | 0 | 3 | 7 | 0 | 3 | 10 | 23 | 106 |
| F-Slide 1 | 0 | 5 | 3 | 0 | 0 | 0 | 48 | 3 | 36 | 93 |
| F-Slide 2 | 0 | 18 | 0 | 0 | 3 | 0 | 15 | 0 | 13 | 48 |
| F-Filter | 46 | 8 | 0 | 0 | 60 | 3 | 0 | 3 | 23 | 141 |

## Discussion

This study is the first to identify the presence of microplastics in tear-replacement solutions in single-use vials, adding to a growing body of evidence on the pervasive nature of microplastics ^4^. Over-the-counter topical tear-replacement solutions were analysed in this pilot study as these were commonly available across international markets and readily accessible without a prescription ^19^. Patients with ocular surface disorders are frequently prescribed topical tear-replacement agents and preservative-free formulations, which are thought to be innocuous, with increased frequency of administration considered useful and often encouraged to supplement either an unhealthy and unstable tear film or improve a poor ocular surface. Frequent use of these topical formulations may therefore inadvertently expose the ocular surface to significant levels of microplastics.

Microplastic particles were identified in all eyedrop samples, with PET, PA and PU being the most commonly identified. This is a concerning observation as microplastics have been associated with multiple adverse health impacts ^7–9^, and may adversely impact patients who rely on regular instillation of eyedrops for ophthalmic diseases. A compromised ocular surface such as in severe dry eye disease, Steven Johnson Syndrome and ocular mucous membrane pemphigoid may also be more susceptible to the negative impacts of microplastics. Owing to its small size, microplastics have a high surface-area-to-volume ratio and bioaccessibility and can exert harmful effects at a cellular level ^20^. These are postulated to occur via three main mechanisms - inflammation and oxidative damage, microbial dysbiosis and toxicological effects from additives and sequestrated chemicals ^16^.

In-vitro studies with human cornea and conjunctival epithelial cell lines have demonstrated the uptake of polystyrene particles with subsequent accumulation around the cell nuclei when they were administered topically ^10^. Cytotoxicity of the PS particles resulted in decreased cell viability and was associated with a reduction in proliferation markers. Exposure of the ocular surface of murine models to 50 nm or 2 μm polystyrene microplastics thrice daily for two to four weeks further demonstrated a clinical reduction in tear secretion, compared to both mice receiving normal saline or controls which received no eyedrops ^10^. Further ex-vivo analysis of tissues revealed reduced size and density of conjunctival goblet cells with an irregular arrangement of lacrimal gland acini in mice treated with microplastics. Congruent to findings in human cornea and conjunctival epithelial cell lines, proliferation-related markers such as Ki-67, p63, and K14 were downregulated, while inflammatory cytokines including IL-1α, IL1-β, and IL-6 were upregulated in murine models exposed with their ocular surface exposed to polystyrene microplastics in a time-dependent fashion ^10^.

Microplastics and their degradation products may also harbour toxic chemicals arising from additives during production, or chemicals absorbed from the environment. A contaminant of concern harboured by plastics are per- and polyfluoroalkyl substances (PFAS) which have been shown to induce immune, cardiovascular, endocrine, and hepatic dysfunction and increase the risk of developing malignancies ^21,22^. Contact lenses have been shown to be a source of microplastic exposure, and a recent study showed that contact lens usage was associated with elevated serum PFAS levels among young adults ^14,23^. While tear homeostasis and the blink mechanism help with regulating tear volume, their role in clearing microplastics and contaminants of emerging concern remains unexplored ^24^. This represents an urgent need to characterise and identify potential ocular exposure of microplastics among ophthalmic therapeutics to prevent long-term adverse effects.

This study employed two different methods to determine the presence of microplastics in commercially available tear-replacement solutions. A range of techniques have been described in the literature to analyse microplastics ^16^. These include non-destructive methods such as transmission electron microscopy, scanning electron microscopy, atomic force microscopy, Raman spectroscopy, atomic force microscopy, and destructive methods such as thermal analysis using pyrolysis or thermal desorption gas chromatography-mass spectrometry (Pyr/TD-GC-MS), and other thermogravimetric analysis (TGA)-based method ^16^. There is no consensus on a standardised approach to processing and analysing samples for microplastics. Variations in counts obtained reflect the differences in technique and limitations of each of the approaches taken. For instance, with sample F, although more than 8000 possible microplastic particles were identified in 1 mL of the sample using light microscopy visualisation, only 48 to 141 particles were definitively identified as microplastics using triplicate analysis with LDIR. This may be related to the constituents of the sample, which contains castor oil and carbomer copolymer type A particles.

This highlights the limitations of visual identification with light microscopy in microplastics analysis. The variation of microplastic counts during visual identification with light microscopy ranges between 20-70 % and increases with smaller particle sizes ^25,26^. Factors that affect identification rates include the operator’s experience, the complexity of the sample matrix, microplastic particle size and the quality and magnification of the microscope ^16,25,27^. Therefore, it is recommended that additional analytical methods be performed in conjunction with light microscopy to reduce error rates and overcome its limitations ^16^.

The LDIR can reliably measure particle sizes from 20 μm in size, and further identify the composition of each individual particle through spectroscopic comparison against established spectral libraries. In the current study, the number of microplastics varied even among the same brands of tear-replacement solutions (Figure 3). This may be related to analytical methodology and environmental confounders which could not be controlled. The Agilent LDIR imaging system possesses the ability to automate the analysis of samples for microplastics. Underestimation may ensue when particles in the sample agglomerate or stack on top of each other, that requires an operator to manually analyse inaccurate components ^28^. The identification capability of LDIR is dependent on the robustness and extensiveness of the spectral library used. Environmental factors such as physical stress, heat, chemical additives, and contamination may alter the morphology and chemical composition of microplastics. These factors are influenced by manufacturing, transport, and storage conditions of the tear-replacement solutions before procurement in this study. Although samples from two production batches were acquired when possible, it is difficult to control and assess the extent of degradation of plastic vials prior to analysis. Moreover, analysis was performed in this study by referencing both a proprietary and in-house spectral database. These databases are not exhaustive and further work remains useful to characterise the spectral characteristics of microplastics to improve the accuracy of current spectroscopic methods ^29–31^. In this study, a spectral match quality of 70% between the sample particle and database was considered to be strong evidence that the particle is of the same polymer type as the reference. Currently, no universally accepted guidelines exist, but previous studies have suggested 70-80% match quality as the cut-off for polymer-type identification ^32,33^. The California State Water Resources Control Board defines 70% or above as the threshold for an accurate spectral match ^34^. Further studies investigating the impact of temperature, physical stress, ultraviolet light, and other environmental factors experienced during the production, transport and storage of tear-replacement solution vials are required to ascertain whether the majority of microplastics arise from manufacturing processes, or from post-production conditions.

Smaller microplastics particles have the propensity to exert more deleterious effects on cells. This is postulated to be related to increased bioaccessibility and altered surface chemistry ^35^. Smaller nanoplastic particles have been reported to cause greater intestinal mucosal damage, while larger nanoplastic particles induce greater microbial dysbiosis in tilapias (*Oreochromis niloticus*) ^36^. In a human intestinal in-vitro model (Caco-2 cell line), smaller 100 nm polystyrene microplastic particles were associated with increased intestinal toxicity compared to 5 μm particles ^37^. As LDIR can reliably identify particles from 20 μm, smaller particles including nanoplastics (<1 nm) were not analysed in the current study. Further characterisation of these particulate properties with techniques such as pyrolysis or thermal desorption gas chromatography-mass spectrometry conferring higher resolution compared to LDIR will be beneficial in future studies.

The impact of microplastics on human health represents an evolving and developing field. Although microplastics were identified from all tested samples, safe levels of exposure and the potential physiological and toxicological effects of microplastics have yet to be determined. This is compounded by the extensive range of chemicals and additives used in the manufacture of different types of plastics. Cradle-to-consumer conditions, including environmental conditions during storage and transport, may influence the stability of plastics housing these drops and contribute to the leeching of chemicals or additives into the topical tear replacement formulations. Future work to quantify microplastics in a range of topical ophthalmic formulations and develop an understanding of inherent risk factors contributing to the instability of the microplastics should be considered. Effects of microplastics retention from chronic instillation over the ocular surface can also be investigated via animal and human studies.

## Conclusion

This study demonstrated the presence of microplastics in commonly used tear-replacement solutions. The discovery of microplastics in therapeutics used on the ocular surface, including contact lenses and eyedrops, raises significant concerns regarding their impact on the ocular surface. Therefore, it is crucial to direct our attention towards identifying, characterising, and addressing potential microplastics throughout the lifecycle of these products. Only by doing so can we take proactive steps to minimise ocular surface exposure to microplastics and reduce potential damage to the ocular surface, especially among patients who rely chronically on these therapeutic agents.

## Data Availability

All data produced in the present work are contained in the manuscript

## Acknowledgements

The study was partly funded by the SERI-Lee Foundation Grant (R1845/87/2021). We thank Eurofins Scientific for supporting this study with experimentation and microplastics analysis with laser direct infrared imaging system with spectroscopy. Illustration was created with BioRender.com.

